# Cost-utility Analysis of Different Treatment Modalities in Patients Undergoing Percutaneous Coronary Intervention: Thai PCI Registry

**DOI:** 10.1101/2024.10.10.24315276

**Authors:** Oraluck Pattanaprateep, Thosaphol Limpijankit, Thunyarat Anothaisintawee, Sukanya Siriyotha, Nakarin Sansanayudh, Ammarin Thakkinstian

## Abstract

**Background:** Cost-utility analyses of percutaneous coronary interventions (PCIs) provide valuable insights into the economic value of these procedures. This analysis aimed to compare the cost-utility of different treatment modalities used for PCI patients within a Thai PCI registry.

**Methods:** Clinical and quality-of-life data from 39 hospitals spread throughout Thailand were used in this analysis. Three treatment modalities for PCI procedures were compared: a) radial vs. femoral access, b) partial vs. complete revascularization for multivessel coronary artery disease, and c) use vs. non-use of device-guided PCIs (intracoronary ultrasound, optical coherence tomography or fractional flow reserve). Cost data were obtained from each patient’s bill for their hospital stay. Utility scores were measured using the Thai EQ-5D-5L at admission, discharge, and one-year post-procedure. The incremental cost-effectiveness ratio (ICER) was then estimated and used for each comparison.

**Results:** Of 19,701 patients enrolled in 2018-19, approximately one-third (n=7,332) had hospital cost data available for analysis. At discharge, the mean costs associated with radial access and partial revascularization were $986 and $391, respectively, less than the costs for device-guided PCIs, which increased by a mean of $2004. The delta utility gains for these approaches were 2.97, 1.60, and 1.07 at discharge and 1.08, 1.06, and 0.29 at one year, respectively. The estimated ICERs at one year were -$894, -$359, and $7041 per unit of utility gained, respectively.

**Conclusion:** Radial access and partial revascularization demonstrated cost-effectiveness, while device-guided PCI incurred high costs for the achieved utility scores. Identifying the factors that influence these outcomes may assist clinicians in tailoring interventions to optimize a patient’s quality of life after PCI.

## Introduction

Percutaneous coronary intervention (PCI), commonly known as coronary angioplasty or stenting, is a procedure performed to open narrowed or blocked coronary arteries. This is done primarily to alleviate symptoms and reduce the risk of myocardial infarction (MI) in patients with coronary artery disease (CAD) (1,2). The global utilization of PCIs has increased over the decades due to improved treatment outcomes; however, the procedures also impose a significant socioeconomic burden (3). As novel PCI techniques and technologies continue to evolve, consideration of economic factors in treatment decisions has become increasingly important (4).

Cost-utility analysis (CUA) enhances transparency in healthcare decision-making by quantifying the trade-offs between costs and utility outcomes (5,6). This approach enables stakeholders to understand the economic implications of various healthcare interventions and holds decision-makers accountable for their resource allocation choices. Three major treatment modalities in PCI that impact CUA are: 1) access site, 2) completeness of revascularization, and 3) the adjunctive use of device-guided PCI incorporating intracoronary imaging or physiological assessment.

Previous studies have shown that transradial intervention is associated with lower costs, more favorable clinical outcomes, and higher patient satisfaction compared to transfemoral intervention ((7–9). This approach has gained popularity, particularly in Asia. It is used in both acute coronary syndrome (ACS) and clinically stable CAD with lower costs and more favorable clinical outcomes relative to transfemoral intervention (10). A debate over the pros and cons of partial versus complete percutaneous coronary revascularization continues. While complete revascularization often leads to better long-term clinical outcomes, it typically incurs higher upfront costs than partial revascularization (11,12). Additionally, patients who undergo multiple PCIs for complete revascularization are at increased risk for in-stent restenosis, in-stent neoatherosclerosis, and stent thrombosis, requiring repeat revascularization (13–15). Furthermore, they potentially experience a higher risk of bleeding due to long-term dual antiplatelet therapy (16).

New device-guided methods include intracoronary imaging, such as intravascular ultrasound study (IVUS) and optical coherence tomography (OCT), and physiological assessments like fractional flow reserve (FFR). Using clinical trials, registries, and meta-analyses, these methods have been shown to reduce the incidence of major adverse cardiovascular events (MACEs), cardiac death, and the need for target-vessel revascularization (TVR) following PCI (17–20). These techniques provide crucial assessments of lesion morphology and hemodynamics, allowing for improved clinical outcomes. However, uncertainty remains regarding whether the upfront costs of these treatment modalities are offset by fewer adverse cardiovascular (CV) events and thus result in greater cost-effectiveness over time (21).

Evaluation of the cost-effectiveness of emerging PCI modalities will generate crucial data for informing interventionists and policymakers in their decisions regarding resource allocations, reimbursement policies, and treatment guidelines. By comprehensively performing CUAs of these treatment modalities, the results can offer valuable insights for interventionists and help optimize patient outcomes while managing costs. Currently, the few CUA studies that have been done on PCIs come predominantly from high-income countries (22–24).

In Thailand, few CUAs have been done for CAD events following PCI (25,26). Our study aimed to address this knowledge gap and ensure the cost-effectiveness of PCI treatment for the Thai population. The specific objectives of this analysis were to compare CUAs of three different treatment modalities for PCI, using data from a Thai PCI registry: a) radial versus femoral access, b) partial versus complete revascularization for multivessel CAD, and c) use versus non-use of device-guidance (IVUS, OCT or FFR) in PCI. These subgroup analyses will help identify the procedures that will provide the most patient and economic benefit from PCI.

## Materials and Methods

This analysis utilized data from the nationwide multicenter Thai PCI Registry, initiated in 2018 by the Cardiac Intervention Association of Thailand. The research methodology has been described in detail elsewhere (27). In brief, data were collected from 39 hospitals, including university, government, and private facilities, across five country regions. Adult subjects (18 years and older) were enrolled in the analysis if they underwent PCI between May 1, 2018, and April 2, 2019, or between June 21 and August 1, 2019, and provided written informed consent. The analysis was approved by the Faculty of Medicine Ethics Committee, Ramathibodi Hospital, Mahidol University (COA-MURA2022/205). Participants were followed for one year after hospital discharge from PCI.

### Interventions of Interest

Three different treatment modalities for PCI procedures were compared: a) radial versus femoral access, b) partial versus complete revascularization for multivessel CAD (including staged PCI within three months), and c) use versus non-use of device-guidance (IVUS, OCT, or FFR) in PCI.

### Data Collection

Demographic, clinical, angiographic, and procedural data were retrieved from the registry’s main electronic databases. Demographic information included age, gender, and reimbursement scheme (i.e., universal health coverage, civil servant medical benefits, social security service, and uninsured or self-pay). Data from physical examinations and underlying diseases encompassed body-mass index (BMI), the presence of cardiovascular risk factors [such as DM, hypertension, dyslipidemia, CKD (defined as eGFR<60 ml/min/1.73m^2^), and smoking], as well as relevant medical history, including cerebrovascular disease, MI, heart failure, and previous PCI or coronary artery bypass graft (CABG).

Clinical and angiographic data included clinical presentation [ST-elevation myocardial infarction (STEMI), non-ST-elevation myocardial infarction (NSTEMI)/unstable angina (UA), and stable CAD], the number and type of diseased vessels [single vessel disease (SVD), double vessel disease (DVD), triple vessel disease (TVD), and left main (LM)], as well as the presence of ostial lesions, bifurcation lesions, and chronic total occlusions (CTOs). Additional parameters included the number of treated lesions (one or more), the use of stents (one, two, or three or more), the number of treated vessels (one, two, or three or more), access site (radial, femoral, or brachial), and lesion severity assessed utilizing intracoronary imaging (IVUS or OCT) or physiological measurements (FFR).

Furthermore, intra- and post-procedural events were recorded; these included procedure success (defined as residual stenosis <20% with stent treatment or <50% with balloon angioplasty alone) and any procedural complications. These complications encompassed a range of outcomes, such as death, MI, stroke, cardiogenic shock, heart failure, newly required dialysis, access site complications (including bleeding, hematoma, pseudoaneurysm, and arteriovenous fistula), major bleeding (including intracranial or retroperitoneal bleeding, or any bleeding that required blood transfusion), endotracheal intubation, cardioversion/defibrillation, and in-hospital CABG.

At the one-year follow-up, clinical outcomes were assessed, focusing on MACEs, which included MI, stroke, TVR, and CV death.

### Outcomes of Interest

This analysis’s two outcomes of interest were cost and utility. The costs quantitated and analyzed were direct medical expenses, such as procedural costs, hospitalization fees, physician fees, and medication costs. This information was obtained from each patient’s billing records during their procedure admission. The costs were then converted to US dollars using the exchange rate from the Bank of Thailand (1 US dollar = 36.87 Baht as of April 19, 2024) (28).

To determine the utility score, the Thai EQ-5D-5L questionnaire (29) was used to collect data at three points: admission prior to the PCI procedure, discharge after PCI, and one-year follow-up. The EQ-5D-5L is a self-reported measure of a patient’s current health across five dimensions: mobility, self-care (e.g., washing or dressing oneself), usual activities (such as work, study, housework tasks, and leisure activities), pain/discomfort, and anxiety/depression. The questionnaire employs five Likert scales, ranging from "no problems" to "unable/extreme problems". Each resulting score profile was converted into a utility score using Thai coefficients multiplied by 100. The utility scores ranged from -28.30 to 100.00, with values of <0, 0, and 100 representing worse than death, death, and perfect health-related quality of life (HRQoL), respectively (29).

In this analysis, the utility score at admission (prior to the PCI procedure) and the utility score at discharge (or at the one-year follow-up) were compared to compute a “utility gain”. For patients who were unconscious or hemodynamically unstable, HRQoL assessments were conducted later when they had stabilized and could conveniently provide information.

### Statistical Analysis

Patient characteristics in each group were described using mean ± SD for continuous data and frequency (with percentage) for categorical data. For the CUA (5), the incremental cost-effectiveness ratio (ICER) was calculated to determine the incremental cost of additional HRQoL gained (30) for each procedural comparison (i.e., radial versus femoral access, partial versus complete revascularization for multivessel CAD, and use versus non-use of device-guidance in the PCI). This was estimated by dividing the incremental costs by the incremental HRQoL (comparing one-year to baseline).

The incremental costs and utility differences for each treatment comparison were estimated using multivariate linear regression analysis, with femoral access, non-use of lesion severity assessment devices, and complete revascularization for multivessel CAD serving as reference groups. Two equations (eq 1 and 2) were constructed as follows:

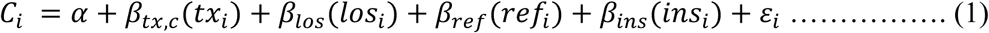

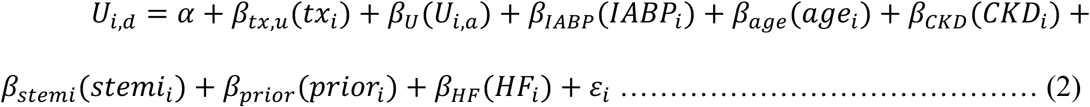

The cost equation (eq 1) was constructed by regressing costs (C_i_) on procedural comparisons (*tx_i_*), length of stay, referral status, and type of health insurance. The utility equation (eq 2) involved regressing utility (U_i_) on procedural comparisons (*tx_i_*), baseline utility score, IABP support, age, CKD, clinical presentation, prior cerebrovascular disease, and heart failure. The ICER was then estimated by dividing *β_tx,c_* with *β_tx,u_*.

Cost-effectiveness ratio planes were plotted from individual patient data for each group comparison to visually illustrate the distribution. All analyses were performed using STATA 18.0 (Stata, TX, USA).

## Results

### Baseline Characteristics and Procedural Details

Out of 19,701 patients enrolled in the registry across 39 PCI centers, more than one-third (n=7,332) had available in-hospital cost data and were included in the analysis. The numbers of subjects available for analyses of the three different procedural modalities were: 7,332 for the comparison of radial versus femoral access, 4,366 for the comparison of partial versus complete revascularization for multivessel CAD, and 7,182 for the use versus non-use of device-guided PCI (IVUS, OCT or FFR), as shown in Figure 1.

**Figure 1.**
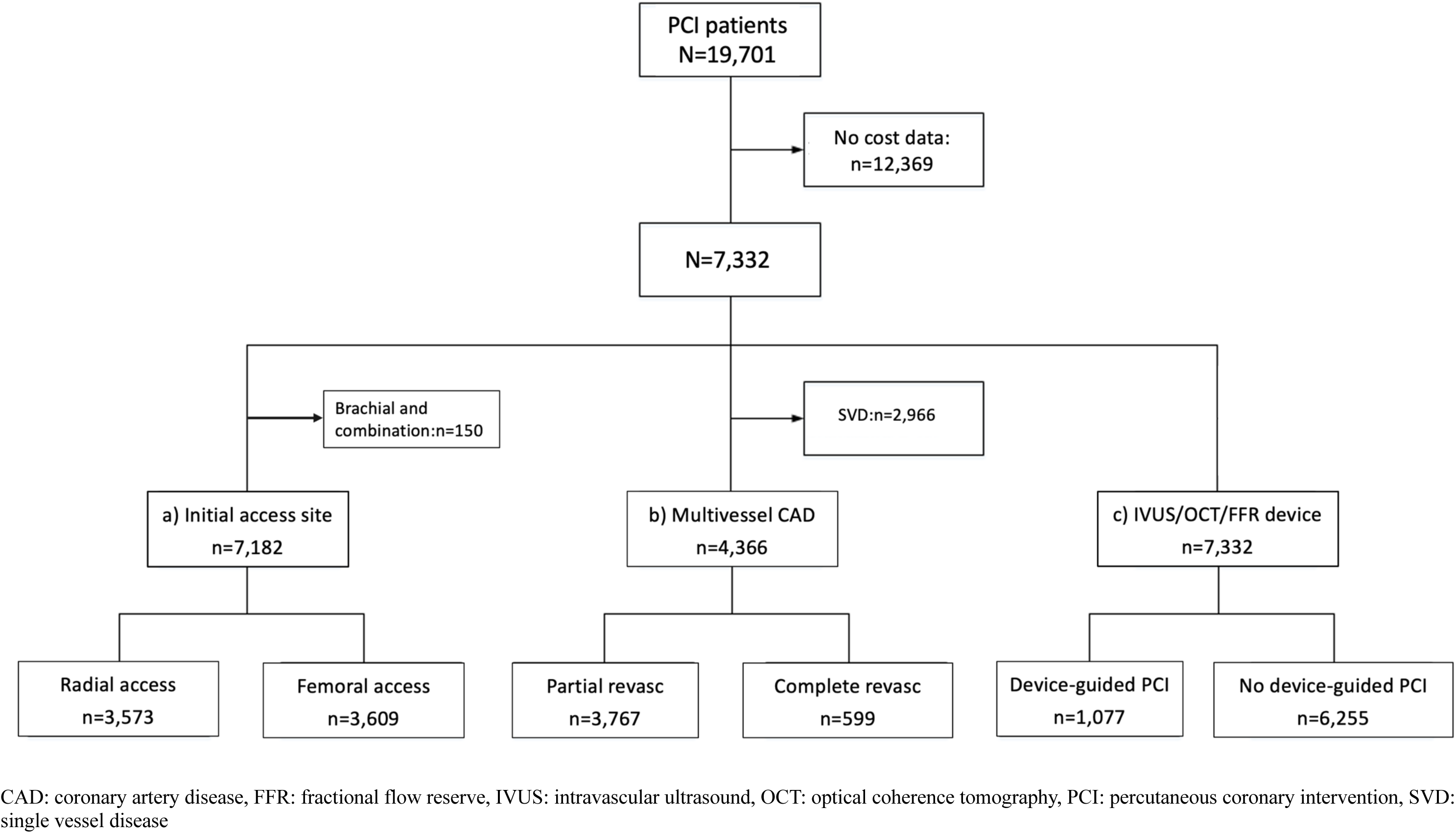
Data flow for three types of categorical treatment modalities during PCI

In the initial comparison of access sites, 3,573 patients (48.7%) were accessed via the radial artery, while 3,609 patients (49.2%) via the femoral artery. One hundred fifty (2.1%) were excluded due to access via the brachial artery or a combination. The comparison of partial and complete revascularization in patients with multivessel disease was comprised of 2,154 with double-vessel CAD and 2,212 with triple-vessel CAD. Among these, 3,767 patients (86.3%) underwent partial revascularization, while 599 (13.7%) had complete revascularization. In the device-guided PCI comparison, 6,255 patients (85.3%) underwent PCI guided by angiography, while the remaining 1,077 (14.7%) utilized at least one lesion severity assessment device.

### Radial versus femoral Access

There were notable differences in patient and angiographic lesion characteristics associated with the access site (Table 1). Radial access was more commonly used in younger patients, males, overweight individuals, and those presenting with NSTEMI/UA or STEMI. Additionally, radial access was more frequently utilized in patients enrolled in the universal health coverage scheme. In contrast, femoral access was used more frequently in patients with comorbid conditions such as DM, HT, DLP, CKD, smoking, prior CVD, and a family history of CAD. It was also used more often in patients with a history of MI, HF, prior PCI, or prior CABG.

**Table 1.** Baseline characteristics of patients using radial versus femoral access.

| Characteristics | Total<br>n=7,182 | Radial access<br>n=3,573 | Femoral access<br>n=3,609 | P-value |
| --- | --- | --- | --- | --- |
| Age groups, years, n (%) |  |  |  |  |
| < 45 | 336 (4.7) | 172 (4.8) | 164 (4.5) | <0.001 |
| 45 – 64 | 3,183 (44.3) | 1,757 (49.2) | 1,426 (39.5) |  |
| 65 – 79 | 2,960 (41.2) | 1,396 (39.1) | 1,564 (43.3) |  |
| ≥ 80 | 703 (9.8) | 248 (6.9) | 455 (12.6) |  |
| Male gender, n (%) | 4,834 (67.3) | 2,521 (70.6) | 2,313 (64.1) | <0.001 |
| BMI ≥ 23 kg/m<sup>2</sup>, n (%) | 4,141 (57.7) | 2,123 (59.4) | 2,018 (55.9) | 0.003 |
| CAD presentation, n (%) |  |  |  |  |
| STEMI | 1,976 (27.5) | 987 (27.6) | 989 (27.4) | <0.001 |
| NSTEMI / Unstable Angina | 2,154 (30.0) | 1,149 (32.2) | 1,005 (27.8) |  |
| Stable CAD | 3,052 (42.5) | 1,437 (40.2) | 1,615 (44.7) |  |
| DM, n (%) | 3,074 (42.8) | 1,439 (40.3) | 1,635 (45.3) | <0.001 |
| HT, n (%) | 4,744 (66.1) | 2,273 (63.6) | 2,471 (68.5) | <0.001 |
| DLP, n (%) | 4,660 (64.9) | 2,254 (63.1) | 2,406 (66.7) | 0.001 |
| Family history of premature CAD, n (%) | 634 (8.8) | 269 (7.5) | 365 (10.1) | <0.001 |
| Cerebrovascular disease, n (%) | 408 (5.7) | 176 (4.9) | 232 (6.4) | 0.006 |
| CKD, n (%) |  |  |  |  |
| With dialysis | 263 (3.7) | 34 (1.0) | 229 (6.3) | <0.001 |
| Without dialysis | 2,241 (31.2) | 927 (25.9) | 1,314 (36.4) |  |
| No | 4,678 (65.1) | 2,612 (73.1) | 2,066 (57.2) |  |
| Current/Ex-smoker, n (%) | 1,542 (21.5) | 850 (23.8) | 692 (19.2) | <0.001 |
| Prior MI, n (%) | 1,989 (27.7) | 932 (26.1) | 1,057 (29.3) | 0.002 |
| Prior heart failure, n (%) | 1,097 (15.3) | 439 (12.3) | 658 (18.2) | <0.001 |
| Prior PCI, n (%) | 2,172 (30.2) | 989 (27.7) | 1,183 (32.8) | <0.001 |
| Prior CABG, n (%) | 112 (1.6) | 8 (0.2) | 104 (2.9) | <0.001 |
| Health insurance, n (%) |  |  |  |  |
| Universal coverage | 4,363 (60.7) | 2,374 (66.4) | 1,989 (55.1) | <0.001 |
| Government service/state enterprise | 2,320 (32.3) | 955 (26.7) | 1,365 (37.8) |  |
| Social security service | 375 (5.2) | 198 (5.5) | 177 (4.9) |  |
| Uninsured or self-pay | 124 (1.7) | 46 (1.3) | 78 (2.2) |  |
| Number of diseased vessels, n (%) |  |  |  |  |
| SVD | 2,107 (29.3) | 1,245 (34.8) | 862 (23.9) | <0.001 |
| DVD | 2,106 (29.3) | 1,075 (30.1) | 1,031 (28.6) |  |
| TVD | 2,171 (30.2) | 969 (27.1) | 1,202 (33.3) |  |
| LM | 798 (11.1) | 284 (8.0) | 514 (14.2) |  |
| Ostial lesion, n (%) | 933 (13.0) | 430 (12.1) | 503 (14.0) | 0.016 |
| Bifurcation, n (%) | 1,237 (17.3) | 512 (14.4) | 725 (20.2) | <0.001 |
| CTO lesion, n (%) | 104 (1.5) | 44 (1.2) | 60 (1.7) | 0.125 |
| Number of treated lesions, n (%) |  |  |  |  |
| 1 | 6,229 (86.7) | 3,110 (87.0) | 3,119 (86.4) | 0.440 |
| > 1 | 953 (13.3) | 463 (13.0) | 490 (13.6) |  |
| Number of stents used, n (%) |  |  |  |  |
| 1 | 3,604 (54.2) | 1,916 (57.2) | 1,688 (51.1) | <0.001 |
| 2 | 2,088 (31.4) | 1,031 (30.8) | 1,057 (32.0) |  |
| ≥ 3 | 963 (14.5) | 404 (12.1) | 559 (16.9) |  |
| Number of treated vessels, n (%) |  |  |  |  |
| 1 | 5,677 (79.0) | 2,864 (80.2) | 2,813 (77.9) | 0.070 |
| 2 | 1,275 (17.8) | 600 (16.8) | 675 (18.7) |  |
| ≥ 3 | 230 (3.2) | 109 (3.1) | 121 (3.4) |  |
| Procedure success, n (%) | 6,833 (95.1) | 3,449 (96.5) | 3,384 (93.8) | <0.001 |
| Procedure complications, n (%) | 451 (6.3) | 176 (4.9) | 275 (7.6) | <0.001 |
| Access site complication, n (%) | 30 (0.4) | 11 (0.3) | 19 (0.5) | 0.151 |
| Major bleeding, n (%) | 90 (1.3) | 16 (0.5) | 74 (2.1) | <0.001 |
| MI, n (%) | 244 (3.4) | 108 (3.0) | 136 (3.8) | 0.081 |
| 1-year MACE, n (%) | 1,076 (15.0) | 408 (11.4) | 668 (18.5) | <0.001 |
| TVR, n (%) | 904 (12.6) | 460 (12.9) | 444 (12.3) | 0.465 |
| MI, n (%) | 346 (4.8) | 150 (4.2) | 196 (5.4) | 0.015 |
| CV death, n (%) | 366 (5.4) | 121 (3.5) | 245 (7.4) | <0.001 |

BMI: body mass index (kg/m<sup>2</sup>), CABG: coronary artery bypass graft, CAD: coronary artery disease, CKD: chronic kidney disease (eGFR< 60 mL/min/1.73m<sup>2</sup>), CTO, chronic total occlusion, CV: cardiovascular, CVD: cerebrovascular disease, DM: diabetes mellitus, DLP: dyslipidemia, DVD: double vessel disease, HT: hypertension, LM: left main, MACE: major adverse cardiovascular event, MI: myocardial infarction, NSTEMI: non-ST elevation MI, PCI: percutaneous coronary intervention, SD: standard deviation, STEMI: ST-elevation myocardial infarction, SVD: single vessel disease, TVD: triple vessel disease, TVR: target vessel revascularization, UA: unstable angina

Regarding lesion characteristics, femoral access was used more frequently in patients with TVD, LM disease, and ostial or bifurcation lesions. However, there were no significant differences based on access in the numbers of lesions and vessels treated, although PCI via femoral access often involved using more than three stents. Radial access was associated with greater procedural success (96.5 vs. 93.8%, p < 0.001) and fewer complications (4.9 vs. 7.6%, p < 0.001). In comparison, patients undergoing femoral access experienced more frequent major bleeding (2.1 vs. 0.5%, p < 0.001) and a trend toward more periprocedural MIs (3.8 vs. 3.0%, p =0.081). At the one-year follow-up, radial access was associated with statistically less frequent occurrences of MI (4.2 vs. 5.4%, p =0.015) and CV death (3.5 vs. 7.4%, p < 0.001). However, there was no difference in TVR frequency (Table 1).

### Partial versus Complete Revascularization

The patients with multivessel CAD were grouped by type of revascularization, as shown in Table 2. Patients who underwent partial revascularization were typically older and had higher frequencies of stable CAD, CKD without dialysis, positive smoking status, and STEMI. Patients covered by social security services generally underwent partial revascularization, while uninsured or self-paying patients unexpectedly underwent complete revascularization.

**Table 2.** Baseline characteristics of patients with partial versus complete revascularization for multivessel CAD.

| Characteristics | Total<br>n=4,366 | Partial<br>revascularization<br>n=3,767 | Complete<br>revascularization<br>n=599 | P-value |
| --- | --- | --- | --- | --- |
| Age groups, years, n (%) |  |  |  |  |
| < 45 | 181 (4.1) | 152 (4.0) | 29 (4.8) | 0.006 |
| 45 – 64 | 1,881 (43.1) | 1,587 (42.1) | 294 (49.1) |  |
| 65 – 79 | 1,872 (42.9) | 1,646 (43.7) | 226 (37.7) |  |
| ≥ 80 | 432 (9.9) | 382 (10.1) | 50 (8.3) |  |
| Male gender, n (%) | 2,934 (67.2) | 2,535 (67.3) | 399 (66.6) | 0.740 |
| BMI ≥ 23 kg/m<sup>2</sup>, n (%) | 2,539 (58.2) | 2,173 (57.7) | 366 (61.1) | 0.120 |
| CAD presentation, n (%) |  |  |  | <0.001 |
| STEMI | 1,005 (23.0) | 897 (23.8) | 108 (18.0) |  |
| NSTEMI / Unstable Angina | 1,217 (27.9) | 976 (25.9) | 241 (40.2) |  |
| Stable CAD | 2,144 (49.1) | 1,894 (50.3) | 250 (41.7) |  |
| DM, n (%) | 1,996 (45.7) | 1,728 (45.9) | 268 (44.7) | 0.610 |
| HT, n (%) | 3,029 (69.4) | 2,602 (69.1) | 427 (71.3) | 0.280 |
| DLP, n (%) | 2,992 (68.5) | 2,593 (68.8) | 399 (66.6) | 0.280 |
| Family history of premature CAD, n (%) | 391 (9.0) | 334 (8.9) | 57 (9.5) | 0.610 |
| Cerebrovascular disease, n (%) | 259 (5.9) | 226 (6.0) | 33 (5.5) | 0.640 |
| CKD, n (%) |  |  |  | 0.016 |
| With dialysis | 176 (4.0) | 146 (3.9) | 30 (5.0) |  |
| Without dialysis | 1,428 (32.7) | 1,261 (33.5) | 167 (27.9) |  |
| No | 2,762 (63.3) | 2,360 (62.6) | 402 (67.1) |  |
| Current/Ex-smoker, n (%) | 869 (19.9) | 770 (20.4) | 99 (16.5) | 0.026 |
| Prior MI, n (%) | 1,466 (33.6) | 1,320 (35.0) | 146 (24.4) | <0.001 |
| Prior heart failure, n (%) | 741 (17.0) | 645 (17.1) | 96 (16.0) | 0.510 |
| Prior PCI, n (%) | 1,722 (39.4) | 1,574 (41.8) | 148 (24.7) | <0.001 |
| Prior CABG, n (%) | 62 (1.4) | 55 (1.5) | 7 (1.2) | 0.580 |
| Health insurance, n (%) |  |  |  | 0.016 |
| Universal coverage | 2,667 (61.1) | 2,307 (61.2) | 360 (60.1) |  |
| Government service/state enterprise | 1,419 (32.5) | 1,224 (32.5) | 195 (32.6) |  |
| Social security service | 208 (4.8) | 183 (4.9) | 25 (4.2) |  |
| Others | 72 (1.6) | 53 (1.4) | 19 (3.2) |  |
| Number of diseased vessels, n (%) |  |  |  | <0.001 |
| DVD | 2,154 (49.3) | 1,669 (44.3) | 485 (81.0) |  |
| TVD | 2,212 (50.7) | 2,098 (55.7) | 114 (14.0) |  |
| Ostial lesion, n (%) | 500 (11.5) | 394 (10.5) | 106 (17.7) | <0.001 |
| Bifurcation, n (%) | 677 (15.6) | 526 (14.0) | 151 (25.2) | <0.001 |
| CTO lesion, n (%) | 61 (1.4) | 51 (1.4) | 10 (1.7) | 0.546 |
| Number of treated lesions, n (%) |  |  |  | <0.001 |
| 1 | 3,587 (82.2) | 3,406 (90.4) | 181 (30.2) |  |
| > 1 | 779 (17.8) | 361 (9.6) | 418 (69.8) |  |
| Number of stents used, n (%) |  |  |  |  |
| 1 | 1,932 (47.7) | 1,882 (54.3) | 50 (8.6) | <0.001 |
| 2 | 1,403 (34.6) | 1,122 (32.4) | 281 (48.1) |  |
| ≥ 3 | 717 (17.7) | 464 (13.4) | 253 (43.3) |  |
| Number of treated vessels, n (%) |  |  |  |  |
| 1 | 3,234 (74.1) | 3,234 (85.9) | 0 (0.0) | <0.001 |
| 2 | 953 (21.8) | 533 (14.1) | 420 (70.1) |  |
| ≥ 3 | 179 (4.1) | 0 (0.0) | 179 (29.9) |  |
| Procedure success, n (%) | 4,125 (94.5) | 3,568 (94.7) | 557 (93.0) | 0.085 |
| Procedure complications, n (%) | 271 (6.2) | 238 (6.3) | 33 (5.5) | 0.450 |
| Access site complication, n (%) | 15 (0.3) | 13 (0.4) | 2 (0.3) | 1.000 |
| Major bleeding, n (%) | 53 (1.2) | 49 (1.3) | 4 (0.7) | 0.189 |
| MI, n (%) | 133 (3.1) | 116 (3.1) | 17 (2.8) | 0.750 |
| 1-year MACE, n (%) | 633 (14.5) | 554 (14.7) | 79 (13.2) | 0.327 |
| TVR, n (%) | 753 (17.3) | 712 (18.9) | 41 (6.8) | <0.001 |
| MI, n (%) | 198 (4.5) | 179 (4.8) | 19 (3.2) | 0.084 |
| CV death, n (%) | 216 (5.3) | 195 (5.5) | 21 (3.8) | 0.090 |

Abbreviation as in Table 1.

Patients who received complete revascularization typically had DVD, ostial lesions, or bifurcation lesions, but not CTO lesions. Unsurprisingly, those undergoing complete revascularization had more treated lesions, treated vessels, and stents than those undergoing partial revascularization. There were no statistically significant differences in frequencies of procedural success nor complications (including access site complications, major bleeding, and periprocedural MI) between partial and complete revascularization. At one-year follow-up, patients who underwent partial revascularization experienced a higher frequency of TVR (18.9 vs. 6.8%, p < 0.001) and showed trends toward increased rates of MI (4.8 vs. 3.2%, p =0.084) and CV death (5.5 vs. 3.8%, p =0.090).

### Use versus Non-use of Device-Guided PCI

The adjunctive use of a lesion-severity assessment device was frequently employed in overweight patients with stable CAD and coexisting comorbidities (such as DM, HT, DLP, CKD, prior MI, prior HF, prior PCI, and prior CABG, excluding smoking), as shown in Table 3. The civil servant medical benefits scheme also typically covered patients who used these devices.

**Table 3.** Baseline characteristics of patients who underwent PCI with or without device (IVUS/OCT/FFR) guidance.

| Characteristics | Total<br>n=7,332 | Device-guided<br>PCI<br>n=1,077 | No Device-guided<br>PCI<br>n=6,255 | P-value |
| --- | --- | --- | --- | --- |
| Age groups, years, n (%) |  |  |  |  |
| < 45 | 347 (4.7) | 52 (4.8) | 295 (4.7) | 0.830 |
| 45 – 64 | 3,250 (44.3) | 468 (43.5) | 2,782 (44.5) |  |
| 65 – 79 | 3,018 (41.2) | 456 (42.3) | 2,562 (41.0) |  |
| ≥ 80 | 717 (9.8) | 101 (9.4) | 616 (9.8) |  |
| Male gender, n (%) | 4,947 (67.5) | 719 (66.8) | 4,228 (67.6) | 0.590 |
| BMI ≥ 23 kg/m<sup>2</sup>, n (%) | 4,233 (57.7) | 654 (60.7) | 3,579 (57.2) | 0.032 |
| CAD presentation, n (%) |  |  |  | <0.001 |
| STEMI | 2,014 (27.5) | 123 (11.4) | 1,891 (30.2) |  |
| NSTEMI / UA | 2,195 (29.9) | 307 (28.5) | 1,888 (30.2) |  |
| Stable CAD | 3,123 (42.6) | 647 (60.1) | 2,476 (39.6) |  |
| DM, n (%) | 3,134 (42.7) | 498 (46.2) | 2,636 (42.1) | 0.012 |
| HT, n (%) | 4,843 (66.1) | 765 (71.0) | 4,078 (65.2) | <0.001 |
| DLP, n (%) | 4,765 (65.0) | 760 (70.6) | 4,005 (64.0) | <0.001 |
| Family history of CAD, n (%) | 642 (8.8) | 122 (11.3) | 520 (8.3) | 0.001 |
| CVD, n (%) | 418 (5.7) | 70 (6.5) | 348 (5.6) | 0.220 |
| CKD, n (%) |  |  |  | <0.001 |
| With dialysis | 265 (3.6) | 64 (5.9) | 201 (3.2) |  |
| Without dialysis | 2,291 (31.2) | 344 (31.9) | 1,947 (31.1) |  |
| No | 4,776 (65.1) | 669 (62.1) | 4,107 (65.7) |  |
| Current/Ex-smoker, n (%) | 1,575 (21.5) | 138 (12.8) | 1,437 (23.0) | <0.001 |
| Prior MI, n (%) | 2,041 (27.8) | 393 (36.5) | 1,648 (26.3) | <0.001 |
| Prior heart failure, n (%) | 1,137 (15.5) | 232 (21.5) | 905 (14.5) | <0.001 |
| Prior PCI, n (%) | 2,221 (30.3) | 459 (42.6) | 1,762 (28.2) | <0.001 |
| Prior CABG, n (%) | 118 (1.6) | 29 (2.7) | 89 (1.4) | 0.002 |
| Health insurance, n (%) |  |  |  | <0.001 |
| Universal coverage | 4,468 (60.9) | 474 (44.0) | 3,994 (63.9) |  |
| Government service/state enterprise | 2,355 (32.1) | 530 (49.2) | 1,825 (29.2) |  |
| Social security service | 383 (5.2) | 58 (5.4) | 325 (5.2) |  |
| Uninsured or self-pay | 126 (1.7) | 15 (1.4) | 111 (1.8) |  |
| Number of diseased vessels, n (%) |  |  |  | <0.001 |
| SVD | 2,135 (29.1) | 228 (21.2) | 1,907 (30.5) |  |
| DVD | 2,154 (29.4) | 264 (24.5) | 1,890 (30.2) |  |
| TVD | 2,212 (30.2) | 345 (32.0) | 1,867 (29.9) |  |
| LM | 831 (11.3) | 240 (22.3) | 591 (9.5) |  |
| Ostial lesion, n (%) | 967 (13.2) | 294 (27.3) | 673 (10.8) | <0.001 |
| Bifurcation, n (%) | 1,267 (17.4) | 365 (34.0) | 902 (14.5) | <0.001 |
| CTO lesion, n (%) | 105 (1.4) | 105 (9.8) | 0 (0.0) | <0.001 |
| Number of treated lesions, n (%) |  |  |  |  |
| 1 | 6,358 (86.7) | 876 (81.3) | 5,482 (87.6) | <0.001 |
| > 1 | 974 (13.3) | 201 (18.7) | 773 (12.4) |  |
| Number of stents used, n (%) |  |  |  |  |
| 1 | 3,657 (53.9) | 396 (39.2) | 3,261 (56.5) | <0.001 |
| 2 | 2,134 (31.4) | 366 (36.2) | 1,768 (30.6) |  |
| ≥ 3 | 995 (14.7) | 249 (24.6) | 746 (12.9) |  |
| Number of treated vessels, n (%) |  |  |  |  |
| 1 | 5,798 (79.1) | 779 (72.3) | 5,019 (80.2) | <0.001 |
| 2 | 1,297 (17.7) | 239 (22.2) | 1,058 (16.9) |  |
| ≥ 3 | 237 (3.2) | 59 (5.5) | 178 (2.8) |  |
| Procedure success, n (%) | 6,969 (95.0) | 1,039 (96.5) | 5,930 (94.8) | 0.020 |
| Procedure complications, n (%) | 467 (6.4) | 92 (8.5) | 375 (6.0) | 0.002 |
| Access site complication, n (%) | 31 (0.4) | 12 (1.1) | 19 (0.3) | 0.001 |
| Major bleeding, n (%) | 90 (1.2) | 18 (1.7) | 72 (1.2) | 0.152 |
| MI, n (%) | 249 (3.4) | 43 (4.0) | 206 (3.3) | 0.242 |
| 1-year MACE, n (%) | 1,102 (15.0) | 154 (14.3) | 948 (15.2) | 0.467 |
| TVR, n (%) | 919 (12.5) | 145 (13.5) | 774 (12.4) | 0.319 |
| MI, n (%) | 353 (4.8) | 60 (5.6) | 293 (4.7) | 0.209 |
| CV death, n (%) | 377 (5.5) | 46 (4.5) | 331 (5.6) | 0.157 |

Abbreviation as in Table 1. FFR: fractional flow reserve, IVUS: intravascular ultrasound study, OCT: optical coherence tomography

Regarding lesion characteristics, the adjunctive use of IVUS, OCT, or FFR was more common in patients with TVD, LM lesions, ostial lesions, bifurcation lesions, and CTOs. Following the use of these devices, patients often underwent treatment for multiple CAD lesions, including in multiple vessels and long lesions that required more than 3 stents. As a result, procedural success rates were higher in this group (96.5 vs. 94.8%, p =0.020); however, complications, particularly access site complications, were also more frequent (1.1 vs. 0.3%, p =0.001). Interestingly, there were no significant differences in major bleeding nor periprocedural MI, and the one-year outcomes (including TVR, MI, and CV death) were comparable.

### Cost-utility Analysis

CUAs were conducted at hospital discharge following PCI and at one-year follow-up. Out of 7,332 patients, only 7,214 were included in the CUA at the one-year mark due to missing data for 118 individuals.

For different access sites, the mean cost for patients using femoral access was $5,554, with an improvement in utility score of 14.46. In comparison, patients using radial access incurred a lower cost of -$986 while experiencing an increase in utility of 2.97, resulting in an ICER of -$331 per unit of utility gained at discharge (see Table 4-a). At the one-year follow-up, the average cost decrease for those utilizing radial access was -$964 compared to femoral access. This resulted in a utility score increase of 1.08, with an ICER of -$894 per unit of utility gained.

**Table 4.** Cost-utility analysis at discharge and one year following PCI procedures.

| | Delta cost<br>(US\$) | Delta utility<br>gain | ICER<br>(US\$ per utility gain) |
| --- | --- | --- | --- |
| a) Radial vs. femoral access |  |  |  |
| - at discharge (n = 7,182) | -986.45 | 2.97 | -331.96 |
| - at 1 year (n = 7,060) | -964.61 | 1.08 | -894.82 |
| b) Partial vs. complete revascularization |  |  |  |
| - at discharge (n = 4,366) | -391.52 | 1.60 | -244.77 |
| - at 1 year (n = 4,301) | -379.99 | 1.06 | -359.83 |
| c) Use vs. non-use of device-guided PCI |  |  |  |
| - at discharge (n = 7,332) | 2,004.18 | 1.07 | 1,878.23 |
| - at 1 year (n = 7,214) | 2,020.60 | 0.29 | 7,041.46 |

ICER: Incremental cost-effectiveness ratio

In patients with multivessel CAD, the average cost for complete revascularization was $5,079, with a utility score improvement of 10.95. Conversely, partial revascularization incurred a lower cost of -$391 but resulted in a higher utility gain of 1.60, leading to an ICER of -$244 per unit of utility gained (see Table 4-b). At the one-year follow-up, the average cost decrease for partial revascularization was -$379, resulting in a utility score increase of 1.06 and an ICER of - $359 per unit of utility gained.

From an in-hospital perspective, the average admission cost for patients undergoing PCI treatment with angiography alone was $4,623, resulting in a utility score improvement of 12.86 at the discharge. After adjusting for confounders, the average cost increase associated with the use of any additional lesion severity assessment device was $2,004, leading to a 1.07 utility unit gain at patient discharge, with an ICER of $1,878 per utility unit (see Table 4-c). At the one-year follow-up, the average cost increase for utilizing any additional device-guided PCI was $2,020, resulting in a utility score gain of 0.29, with an ICER of $7,041 per utility unit gained.

### Incremental Cost-effectiveness Ratio (ICER) Analysis

In the ICER analysis at the one-year follow-up, all three comparisons—different access sites, types of revascularizations, and the use of device-guided PCI—showed narrow delta utility gains. The delta utility gains for these approaches were 2.97, 1.60, and 1.07 at discharge and 1.08, 0.06, and 0.29 at the one-year follow-up, respectively (see Table 4). Regarding costs, adding lesion severity assessment devices to angiography resulted in a slight increase of +$16. In contrast, the cost deltas for radial access and partial revascularization were slightly lower (with deltas of -$22 and -$11, respectively).

Consequently, the estimated ICERs were -$894 for radial versus femoral access, -$359 for partial versus complete revascularization, and $7,041 for using severity assessment device-guided PCI versus non-use. Radial access and partial revascularization were deemed cost-effective, as they incurred lower costs with higher effectiveness. In contrast, while device-guided PCI was more expensive, it proved to be more utility than non-use. However, this approach incurs significant costs in obtaining the utility benefit.

The scatter plots of the ICER plane for each comparison at discharge and the one-year follow-up are presented in Figure 2. Overall, the clusters of points indicate that radial access and partial revascularization were cost-effective options, whereas the use of severity assessment device-guided PCI was not. The robustness of the data was consistent between discharge and the one-year follow-up for each comparison.

**Figure 2.**
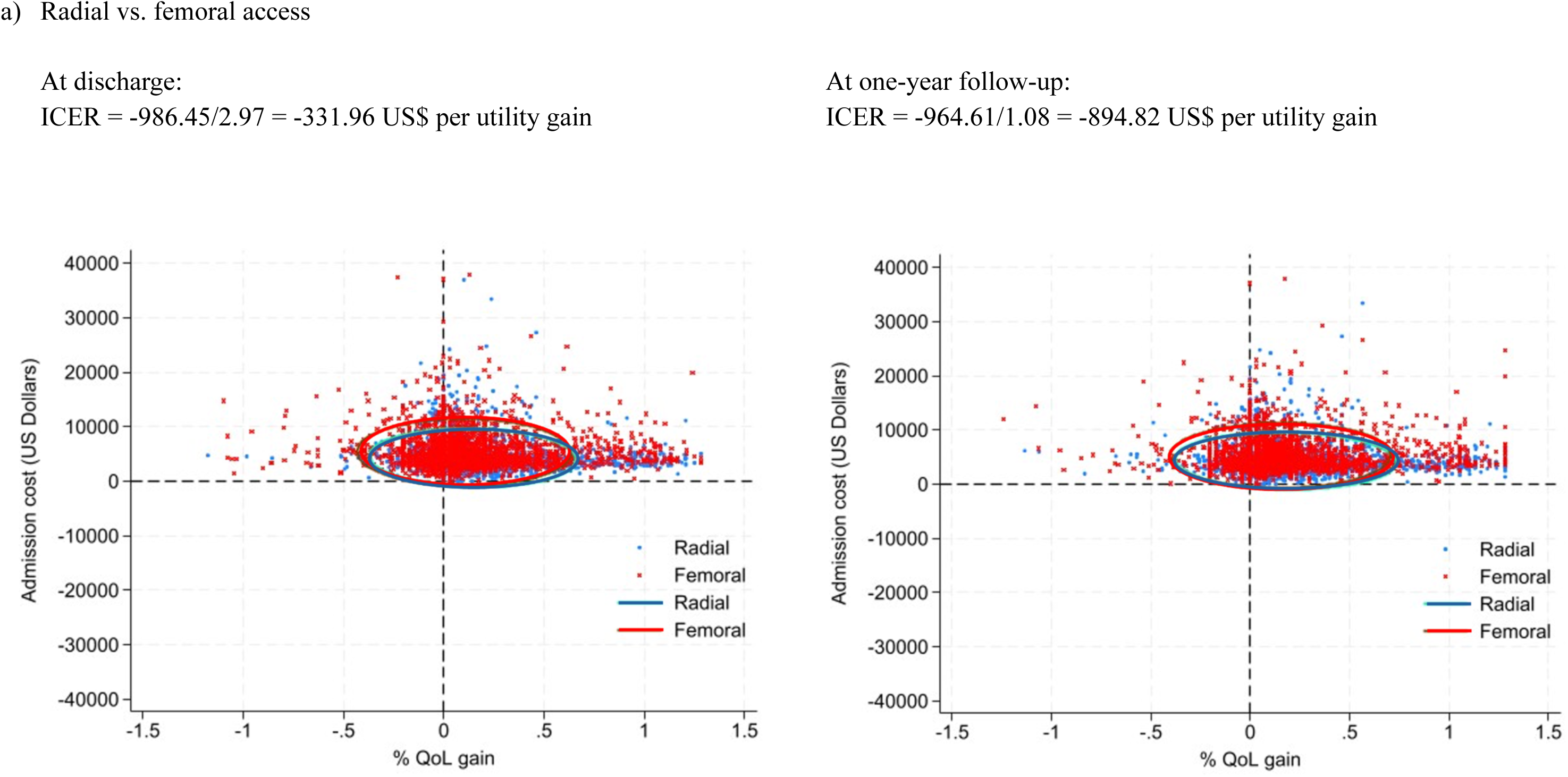
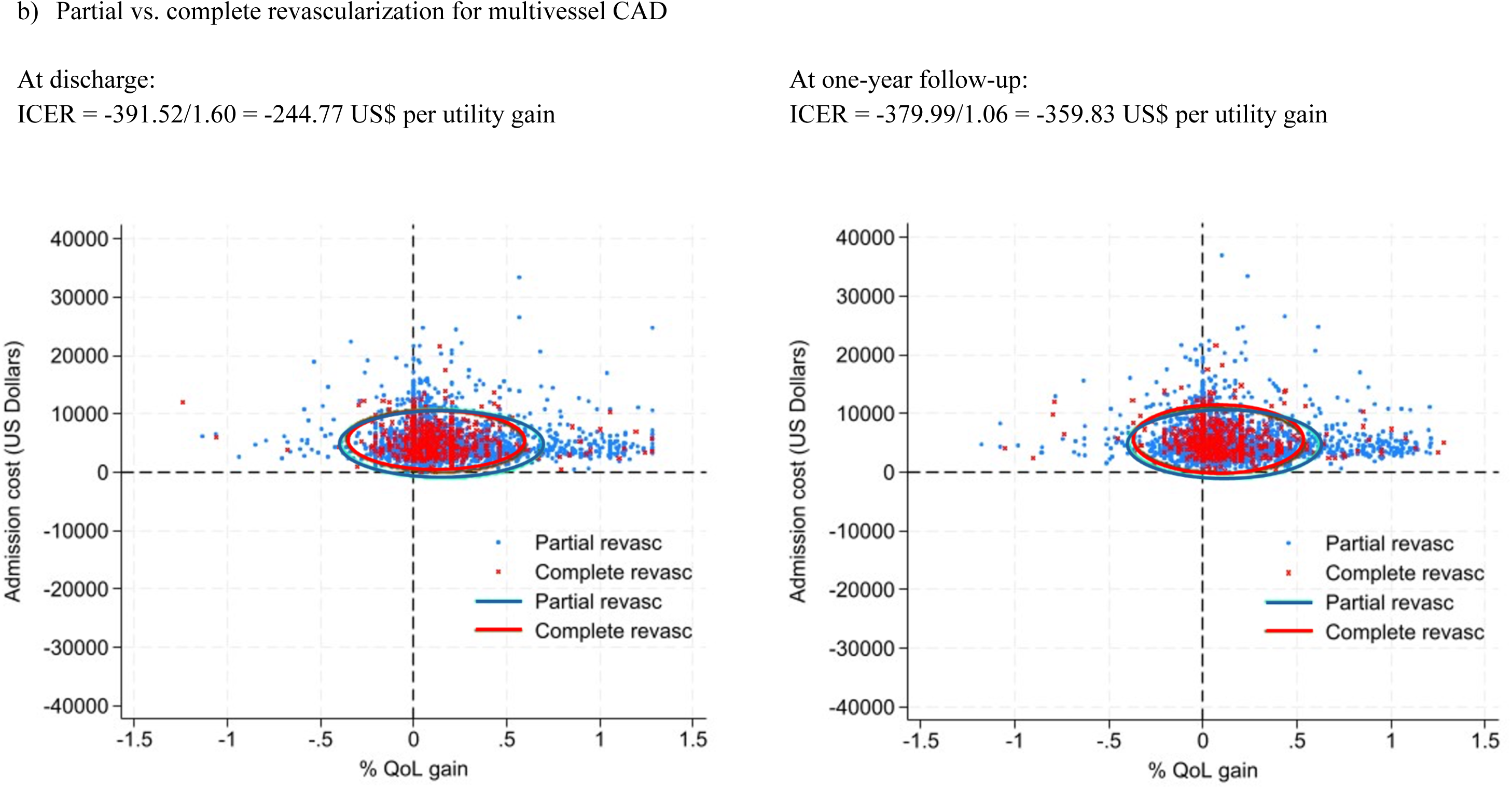
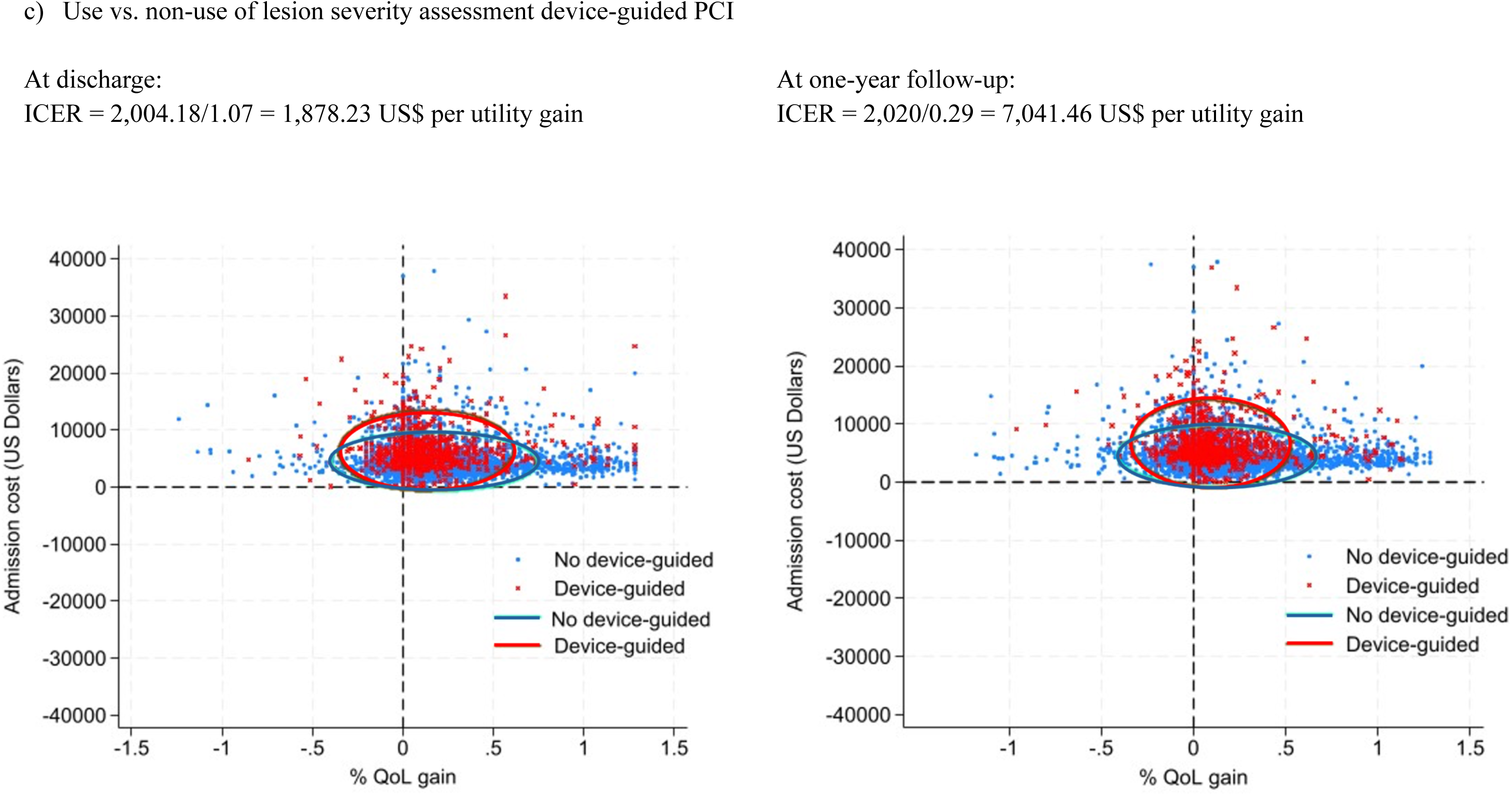
The scatter plots of the incremental cost-effective ratio (ICER) plane for each comparison at discharge and the one-year follow-up.

## Discussion

This CUA utilized data extracted from a nationwide PCI registry to evaluate three different features of the PCI procedures used in Thailand. Our findings highlighted the effectiveness of specific approaches. Notably, adopting a radial access approach and implementing partial revascularization in patients with multivessel CAD emerged as financially advantageous options in both the short (at discharge) and long (after 1 year) term. In contrast, adjunctive lesion severity assessment devices, such as intracoronary imaging and hemodynamic measurements, incurred substantial additional costs with very little improvement in utility.

In contemporary practice, access to the coronary arteries during PCI is achieved via either femoral or radial arteries. Although femoral access has traditionally been favored for its ease of canulation and direct access, substantial evidence supports the benefits of radial access. Radial access is associated with both fewer major bleeds and access site complications compared to femoral access, potentially leading to improved clinical outcomes, shorter hospital stays, and reduced healthcare costs (7,8,22). Additionally, radial access decreases discomfort and accelerates recovery, enhancing patient quality of life and satisfaction (31). Our analysis supports the utilization of radial access due to its cost-utility gains, particularly at discharge but also after one year. This approach should be more cost-effective from a healthcare system perspective, given its potential to reduce overall healthcare expenditures and improve patient outcomes.

In Thailand, the use of radial access for PCI has surpassed femoral access by approximately half, although its adoption still varies. Patient factors, operator preference, and experience all contribute to this variability. Younger interventionists are generally more accustomed to using radial access. Patient factors in our registry include younger age, male gender, overweight status, smoking, fewer comorbidities, presentation with ACS, and universal healthcare coverage. At the one-year follow-up, radial access was associated with lower rates of MACE, particularly MI and CV death. Our findings are consistent with previous studies indicating that radial access correlates with higher procedural success and lower complication rates (7,9). However, it is important to note that operators tend to prefer radial access under stable hemodynamic conditions. In contrast, femoral access is preferred in cases of hemodynamic instability or lesion complexity (e.g., LM or bifurcation lesions) requiring large arterial sheaths or IABP support. Consequently, femoral access is associated with higher costs and lower utility gains, but the subject groups were significantly different. Thus, subgroup analyses could assess the cost-effectiveness of radial versus femoral access for specific patient groups and aid in personalizing treatment.

Regarding revascularization strategy, complete compared to partial revascularization has been associated with better clinical outcomes and improved quality of life, as it alleviates anginal symptoms (11). While complete revascularization usually involves higher upfront procedural costs due to treating additional lesions, it can lead to fewer repeat procedures and hospitalizations, resulting in cost savings over the long term compared to partial revascularization (32–34).

In our study, complete revascularization was performed in 13.7% of patients with multivessel CAD. Partial revascularization was more common, especially among older patients, octogenarians, those with stable CAD, CKD without dialysis, smokers, and individuals without insurance or who self-pay. However, complete revascularization was more frequent in patients presenting with NSTEMI/UA and those with DVD, ostial lesions, and bifurcation lesions (though not in cases of CTO lesions). This resulted in a greater number of stent placements.

Importantly, our study found no difference in procedural success frequencies and complications occurring following partial and complete revascularization. Partial revascularization was associated with lower costs, a crucial consideration for informed resource allocation within healthcare systems. However, the two subject groups differed in underlying disease, and those undergoing partial revascularization exhibited a higher frequency of post-PCI TVR and trends toward more frequent MIs and CV deaths during the one-year follow-up.

This study found that partial revascularization was sufficient, achieving utility scores comparable to those of complete revascularization. This approach conserved healthcare resources while benefiting patients. However, it was essential to carefully select the revascularization strategy based on each patient’s clinical presentation to prevent recurrent MI, TVR, or CV death, especially in a resource-limited setting. For instance, in patients with STEMI or NSTEMI and multivessel CAD undergoing primary PCI of the culprit artery, preventive PCI in non-culprit coronary arteries may be considered if significant residual myocardial ischemia is evident. A mismatch between the visually estimated significance of angiographic coronary stenosis and FFR is common, as visual estimation alone often does not accurately predict functional significance (35). Utilizing FFR measurements concurrently during cardiac catheterization allows for hemodynamic evaluation and can avoid the treatment of hemodynamically non-significant lesions (24,36).

Cost-effectiveness analyses for stable CAD have not favored PCI over medical therapy (6,37,38). Certain patient factors—such as being octogenarians, bedridden with limited activity, experiencing severe renal impairment without planned dialysis, or having secondary NSTEMI— may render patients unsuitable for complete revascularization. Similarly, for CTO lesions with abundant collateral blood supply in asymptomatic or mildly symptomatic patients, revascularization may not be necessary. However, when significant symptoms persist despite medical treatment, complete revascularization may lead to a more cost-efficient outcome (39). However, these conclusions require confirmation through randomized controlled trials. Importantly, CABG remains the gold standard and is more cost-effective than PCI for complex multivessel coronary lesions or LM disease in the long term (12).

The adjunctive use of guided devices, such as IVUS, OCT, and FFR, is associated with improved clinical outcomes compared to PCIs guided by angiography alone. These technologies provide additional information on lesion characteristics, vessel morphology, and hemodynamic significance, optimizing stent placement and enhancing post-PCI outcomes (23,24,40,41). Despite the advances in these technologies and accumulating evidence of their clinical efficacy and safety, their adoption remains limited (42). In our study, the utilization rate was 14.7%, comparable to rates in Europe and the United States (8.7% to 16.5%) (43–45). In contrast, Japan reports much higher utilization rates, ranging from 82% to 91% (46). Key barriers to adoption include prohibitive costs and increased procedure times. Our study found that using these adjunctive devices incurred the highest costs among the three treatment modalities. Furthermore, many interventionists may lack confidence in the technical aspects of advanced intracoronary imaging and its interpretation and be skeptical regarding the cost-effective data supporting their use. Additionally, universal health coverage programs may restrict the adoption of these devices, as they increase treatment costs without significantly reducing adverse events.

Our study revealed that guided devices were predominantly used in cases of stable CAD with complex anatomy and high comorbidities, including CKD, smoking history, prior PCI or CABG, often under government insurance coverage. Adjunctive use was also common in patients with high-risk anatomies, such as TVD, as well as LM, ostial, bifurcation, and CTO lesions. This led to the treatment of numerous lesions and vessels, resulting in increased treatment costs. In terms of outcomes, these devices only marginally improved procedural success but were associated with increased complication rates. Furthermore, our CUA did not support using these guided devices for short-nor long-term (up to 1 year) outcomes, with no significant differences in the incidences of TVR, MI, or CV death. These findings contrast with those of previous studies (23,40) and raise concerns that our cost-benefit analysis did not fully long-term financial benefits. Among Thai interventionists, the younger were more likely to have been trained to use and interpret these guided devices effectively. Subgroup analyses are essential for evaluating the cost-effectiveness of IVUS, OCT, or FFR in specific patient populations, including those with complex lesions, multivessel disease, or high-risk features (47–49). In the near future, intravascular imaging-derived FFR will facilitate a comprehensive assessment of lesions’ anatomical and physiological properties using a single diagnostic device to guide PCI optimization (50).

The CUA of the PCI procedure underscores the importance of evaluating both clinical effectiveness and economic implications, thereby informing decision-making by clinicians, policymakers, and healthcare insurers. Our results can particularly inform healthcare policies in countries with similar economic conditions, potentially improving CV care in many countries. Randomized controlled trials are essential to further clarify the benefits and limitations of each treatment strategy.

## Limitations

A notable strength of this CUA was its access to a national PCI registry, which provided valuable real-world data on the economic implications of PCI for patients with various manifestations of CAD. However, several limitations should be considered when interpreting the findings. First, although data were gathered from 39 out of 72 PCI centers in Thailand, this represents only 54.2 % of the total centers. However, the subjects included in our analysis underwent PCI across a diverse range of catheterization laboratory facilities and from five distinct regions of the country. This geographic and facility diversity strengthens the generalizability of our findings, as it captures a broad spectrum of patient experiences and outcomes. Second, the study utilized solely the hospital bills to patients which may have been incomplete, introducing uncertainty into the economic analysis. Addressing these limitations through sensitivity analyses and validation studies could enhance the reliability of future cost-utility analyses based on similar national registries.

## Conclusion

Our CUA findings suggested that radial access and partial revascularization are cost-effective PCI strategies. However, using lesion severity assessment devices incurred high costs for each unit of utility gained. Further identification of the factors that influence these PCI outcomes should enable clinicians to better tailor interventions to optimize patient quality of life following the procedure.

## Acknowledgments

The study team thanks Dr. Arthur Brown for reviewing the manuscript and providing valuable comments.

## Authors’ contributions

O.P. conceptualization, methodology, study design, data analysis, interpretation, drafting the manuscript, study supervision. T.L. conceptualization, methodology, study design, data analysis, interpretation, writing the manuscript, study supervision. S.S. methodology, data acquisition, data analysis, interpretation. N.S. data acquisition. A.T. conception and design, data analysis, and interpretation of the data, and revision of article and supervision. T.A. conception and design, data analysis, and interpretation of data, and revision of article and supervision. All authors read and approved the final manuscript.

## Data availability statement

The data supporting the findings of this study are available from the corresponding author upon reasonable request.

